# Association of Sleep Behaviors with Risk of Esophageal Cancer

**DOI:** 10.1101/2022.01.23.22269728

**Authors:** Xiaoyan Wang, Ruiyi Tian, Xiaoyu Zong, Myung Sik Jeon, Jingqin Luo, Graham A. Colditz, Jean Wang, Konstantinos K. Tsilidis, Yo-El S Ju, Ramaswamy Govindan, Varun Puri, Yin Cao

**Affiliations:** Division of Public Health Sciences, Department of Surgery, Washington University School of Medicine, St. Louis, USA; Brown School, Washington University in St. Louis, St. Louis, USA; Alvin J. Siteman Cancer Center, Washington University School of Medicine, St. Louis, USA; Division of Gastroenterology, Department of Medicine, Washington University School of Medicine, St. Louis, USA; Department of Epidemiology and Biostatistics, School of Public Health, Imperial College London, London, UK; Department of Hygiene and Epidemiology, University of Ioannina School of Medicine, Ioannina, Greece; Center on Biological Rhythms and Sleep (COBRAS), Washington University School of Medicine, St. Louis, USA; Hope Center for Neurological Disorders, Washington University School of Medicine, St. Louis, USA; Department of Anesthesiology, Washington University School of Medicine, St. Louis, USA; Division of Oncology, Department of Medicine, Washington University School of Medicine, St. Louis, USA; Division of Cardiothoracic Surgery, Department of Surgery, Washington University School of Medicine, St. Louis, USA

## Abstract

**IMPORTANCE:** Esophageal cancer is among the most lethal type of cancers worldwide. However, risk factors contributing to more than tenfold increase in esophageal cancer in the last 50 years remain underexplored.

**OBJECTIVE:** This study aimed to examine the associations between sleep behaviors and esophageal cancer overall, by histology, and according to genetic predispositions.

**DESIGN:** A prospective cohort study.

**SETTING:** A population-based study.

**PARTICIPANTS:** A total of 410,428 participants aged 37-73 years at enrollment between 2006 and 2010 in the UK Biobank were followed up until March 31st, 2016 for England and Wales and October 31st, 2015 for Scotland.

**MAIN OUTCOME AND MEASURE:** The risk of incident esophageal cancer.

**RESULTS:** During 2,799,342 person-years of follow-up, 410 incident esophageal cancer cases (294 adenocarcinomas) were diagnosed. Evening chronotype, sleep <6 or >9 h/day, daytime napping, and daytime sleepiness were significantly associated with increased risk of esophageal cancer in age-adjusted models and had a *P*_*likelihood ratio test*_ ≤0.20 after multivariable adjustment. Compared with the group without these high-risk behaviors, participants with one high-risk behavior had a 41% (HR=1.41, 95%CI: 1.13, 1.77) increased risk of esophageal cancer, and those with two or more behaviors showed a 79% higher risk (HR=1.79, 95%CI: 1.32, 2.42) (*P*_*trend*_<0.001). These associations were largely driven by esophageal adenocarcinoma (*P*_*trend*_<0.001) but not squamous cell carcinoma (*P*_*trend*_=0.340). The elevated risks for esophageal adenocarcinoma were similar within strata of PRS quintiles (*P*_*interaction*_=0.791).

**CONCLUSION AND RELEVANCE:** Unhealthy sleep behaviors were associated with an increased risk of esophageal cancer, primarily adenocarcinoma, independent of genetic risk. Sleep behaviors may serve as modifiable factors for the prevention of esophageal cancer, particularly esophageal adenocarcinomas.

**Key Points:** *Question:* Are sleep behaviors associated with the risk of esophageal cancer?

*Findings:* In this prospective cohort study that included 410,428 participants in the UK Biobank, evening chronotype, sleep <6 or >9 h/day, daytime napping, and sleepiness were associated with increased risk of esophageal cancer. A greater number of these unhealthy sleep behaviors was associated with a higher risk of esophageal cancer. The elevated risks were primarily observed for esophageal adenocarcinoma and were independent of genetic risk.

*Meaning:* Sleep behaviors may serve as modifiable factors for the esophageal cancer prevention, particularly esophageal adenocarcinoma, independent of genetic risk.

## Introduction

Esophageal cancer is among the most lethal type of cancers worldwide, with 5-year overall survival of 15% to 25%.^1^ In western countries, incidence of esophageal adenocarcinoma (EAC), the most common type of esophageal cancer, has been increasing substantially since the 1960s from 0.41 cases to 5.31 cases per 100 000 person-years in 2007,^2^ surpassing esophageal squamous cell carcinoma (ESCC).^2^ However, besides obesity, the contributors to the dramatic increase in EAC have not been identified.^3,4^

Sleep-related behaviors, such as excessively short/long sleep duration and insomnia, have become increasingly prevalent and emerged as a public health epidemic.^5,6^ From 1985 to 2012, US adults sleeping ≤ 6 hours increased from 38.6 to 70.1 million, and up to 38% of individuals aged 25-44 reported sleep duration less than 7 hours.^5^ A meta-analysis from the Netherlands, UK, and US revealed that poor sleep quality (13%) and insomnia symptoms (9.6-19%) were even more prevalent than short sleep duration.^6^ Accumulating evidence supports the link between sleep behaviors and esophageal cancer. For instance, individuals who sleep 5-6 hours have increased risks for Barrett’s esophagus^7^ and ESCC^8^ compared with those who sleep 7-8 hours. Snoring, a symptom of obstructive sleep apnea, was also associated with increased risk for Barrett’s esophagus^9^ and ESCC.^8^ However, other sleep behaviors, such as chronotype, daytime sleepiness, and insomnia, have not yet been examined with risk of esophageal cancer, especially EAC. As these sleep behaviors are usually correlated, there is an imperative need to evaluate the collective impact of multiple sleep behaviors on esophageal cancer risk.^10^

The development of esophageal cancer is in part attributed to genetic factors.^11^ Thus far, a total of 18 single nucleotide polymorphisms (SNPs) have been identified from genome-wide association studies (GWAS) for EAC.^12–14^ Among these SNPs, rs62423175 in MTRNR2L9 was found to be associated with chronotype, duration, daytime napping, daytime sleepiness, snoring, and insomnia,^15^ providing additional rationale to examine the role of sleep disturbance in EAC in the context of genetic predisposition.

To address these knowledge gaps, we prospectively examined the associations between major sleep behaviors (chronotype, duration, daytime napping, daytime sleepiness, snoring, and insomnia) and risk of esophageal cancer overall, by histology, and according to genetic predispositions, leveraging data from the UK Biobank, a large, prospective cohort with a comprehensive assessment of sleep behaviors and genetic profiling.

## Methods

### Study population

The UK Biobank is a population-based prospective study with over half a million participants aged 37-73 years recruited between 2006 and 2010. The detailed study design and methods have been described previously.^16^ In brief, at baseline visit, participants completed a self-administered touchscreen questionnaire and underwent physical examination for collecting sleep and other health-related information, and provided blood for genotyping. The UK Biobank study was approved by the National Information Governance Board for Health and Social Care in England and Wales, the North West Multicentre Research Ethics Committee, and the Community Health Index Advisory Group in Scotland.

Initially, data were obtained from 502,486 participants. After excluding individuals with cancer except for non-melanoma skin cancer prior to the baseline visit or missing values on any of the sleep behaviors, a total of 410,428 participants were included in the analyses (**eFigure 1** in the Supplement).

**Figure 1.**
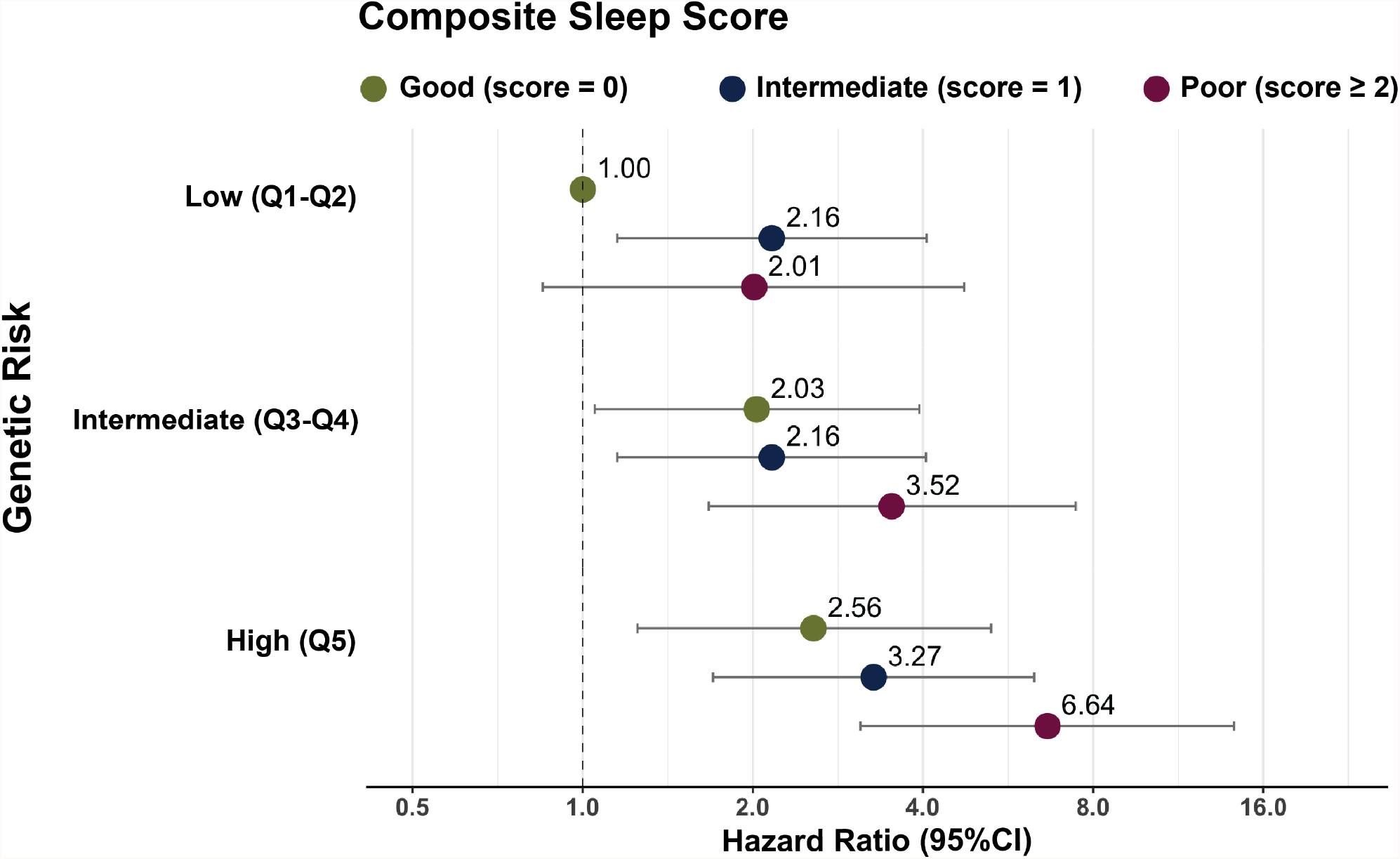
Joint associations of composite sleep score and genetic predisposition with risk of esophageal adenocarcinoma. Composite sleep score was calculated by summing the individual scores of four unhealthy sleep behaviors: evening chronotype, sleep <6 or >9 h/day, daytime napping and usual daytime sleepiness. Each unhealthy sleep behavior was assigned with score 1 otherwise with score 0. The composite sleep score was further categorized into three groups: good (score=0), intermediate (score=1), and poor (score≥2). Genetic risk was defined by quintiles of PRS: low-(Q1-Q2), intermediate-(Q3-Q4), and high-risk (Q5). All hazard ratios were adjusted for the same set of covariates as model b in Table 2, the first ten principal components for ancestry, and the genotype array type. Abbreviation: CI: confidence interval.

### Ascertainment of outcome

Incident esophageal cancer cases were identified through linkage to cancer registries and death records provided by the National Health Service (NHS) Information Centre and the NHS Central Register, National Records of Scotland, and defined using the International Classification of Diseases, Tenth Revision, Clinical Modification (ICD-10-CM) code C15. We further classified esophageal cancer into EAC and ESCC, using the International Classification of Diseases for Oncology, Third Edition (ICD-O-3) codes 8140-8573 for EAC and 8050-8082 for ESCC. Complete follow-up was available up to March 31^st^, 2016 for England and Wales and October 31^st^, 2015 for Scotland.

### Assessment of sleep behaviors

We extracted information on the following six sleep behaviors self-reported through a questionnaire at baseline: chronotype, duration, daytime napping, daytime sleepiness, snoring, and insomnia. Information on chronotype was collected using the question: “Do you consider yourself to be: 1) definitely a “morning” person, 2) more a “morning” than “evening” person, 3) more an “evening” than “morning” person, or 4) definitely an “evening” person” and categorized as morning, more morning than evening, more evening than morning, and evening. Sleep duration was collected from the question: “About how many hours sleep do you get in every 24h? (please include naps)” and collapsed as <6, 6, 7, 8, 9, and >9 h/day. Frequencies of daytime napping were asked through the question: “Do you have a nap during the day?” with responses of never/rarely, sometimes, and usually. Daytime sleepiness was obtained from the question “How likely are you to doze off or fall asleep during the daytime when you don’t mean to? (e.g., when working, reading or driving)” and regrouped as never/rarely, sometimes, and usually (often and all of the time). Snoring was collected by asking “Does your partner or a close relative or friend complain about your snoring?” and grouped as yes and no. Insomnia coded the responses from the question “Do you have trouble falling asleep at night or do you wake up in the middle of the night?” as never/rarely, sometimes, and usually. Prior mendelian randomization studies suggest that the associations between the identified gene loci and self-reported measurement for chronotype,^17^ duration,^18^ daytime napping,^19^ daytime sleepiness,^20^ and insomnia^21^ in the UK Biobank were consistent with those identified using accelerometers.

### Derivation of polygenic risk score

We derived a polygenic risk score (PRS), based on 17 of the 18 SNPs previously identified GWAS-significant SNPs of EAC (*P* < 5 × 10^−8^),^12–14^ to measure participants’ genetic susceptibility to EAC. SNP rs66725070 was not available in the UK Biobank (**eTable 1** in the Supplement). Each SNP was coded based on the number of risk alleles as 0, 1 and 2. Weights were derived based on the reported effect size (log odds) from prior GWASs.^12–14^ For each participant, PRS was calculated as the sum of the product of the SNP and its corresponding weight (**eTable 1** in the Supplement).^22^ We then grouped participants into three groups based on PRS quintiles: low-(Q1-Q2), intermediate-(Q3-Q4), and high-risk (Q5). A total of 278,893 participants were included in the calculation after the exclusions (**eMethods** in the Supplement).

**Table 1.**
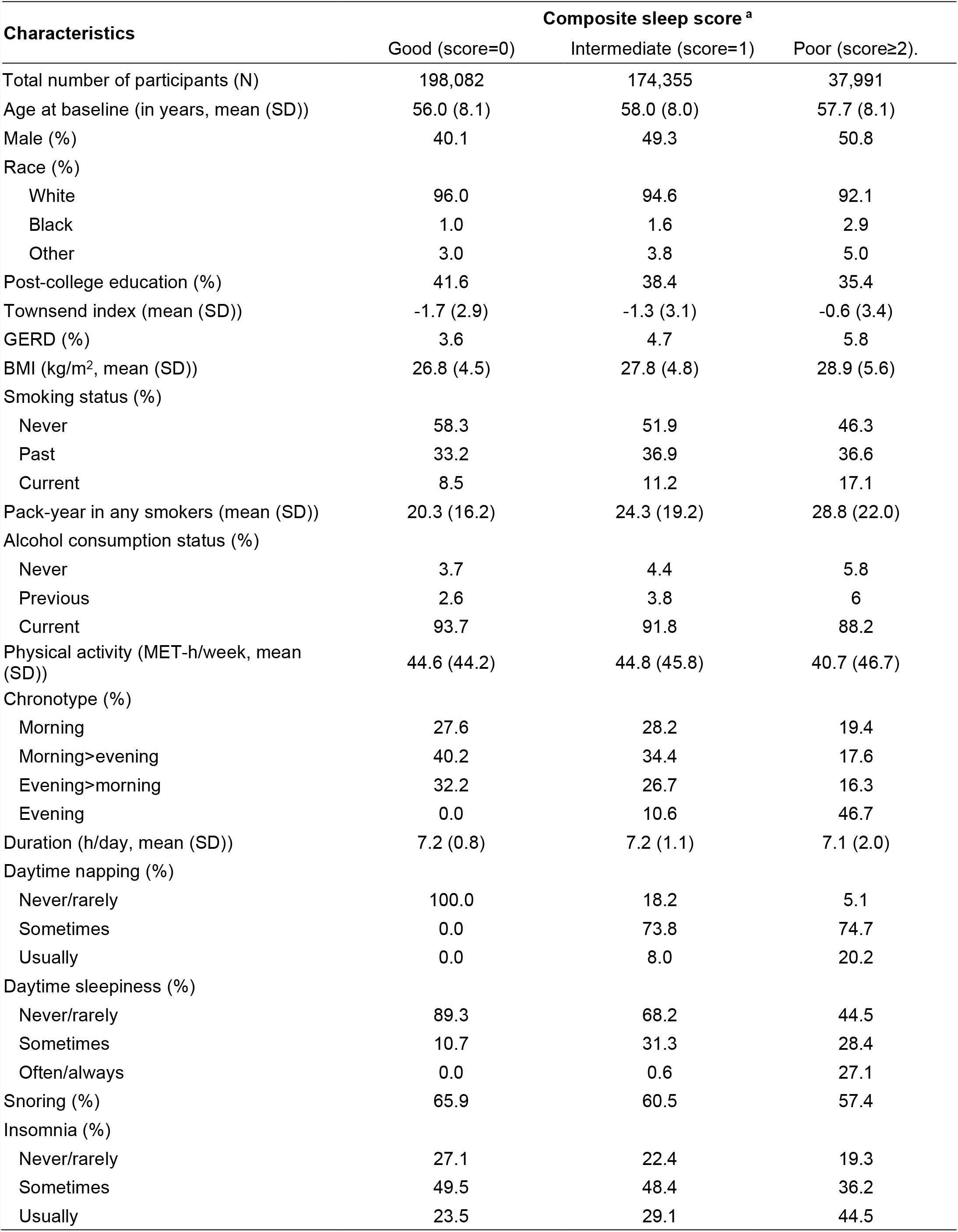
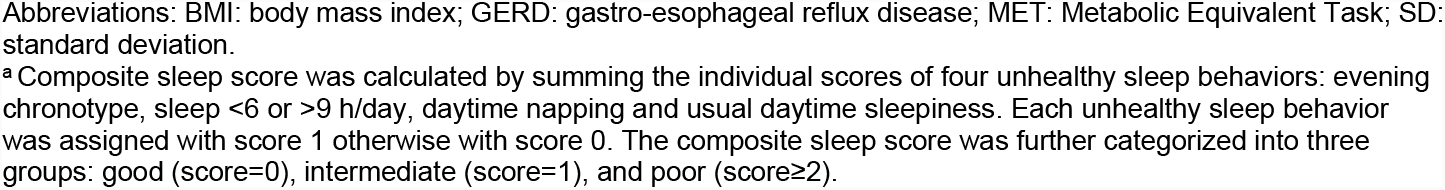
Baseline characteristics of 410,428 participants according to composite sleep score, UK Biobank 2006-2010.

| Characteristics | Composite sleep score <sup>a</sup> |  |  |
| --- | --- | --- | --- |
|  | Good (score=0) | Intermediate (score=1) | Poor (score≥2). |
| Total number of participants (N) | 198,082 | 174,355 | 37,991 |
| Age at baseline (in years, mean (SD)) | 56.0 (8.1) | 58.0 (8.0) | 57.7 (8.1) |
| Male (%) | 40.1 | 49.3 | 50.8 |
| Race (%) |  |  |  |
| White | 96.0 | 94.6 | 92.1 |
| Black | 1.0 | 1.6 | 2.9 |
| Other | 3.0 | 3.8 | 5.0 |
| Post-college education (%) | 41.6 | 38.4 | 35.4 |
| Townsend index (mean (SD)) | -1.7 (2.9) | -1.3 (3.1) | -0.6 (3.4) |
| GERD (%) | 3.6 | 4.7 | 5.8 |
| BMI (kg/m<sup>2</sup>, mean (SD)) | 26.8 (4.5) | 27.8 (4.8) | 28.9 (5.6) |
| Smoking status (%) |  |  |  |
| Never | 58.3 | 51.9 | 46.3 |
| Past | 33.2 | 36.9 | 36.6 |
| Current | 8.5 | 11.2 | 17.1 |
| Pack-year in any smokers (mean (SD)) | 20.3 (16.2) | 24.3 (19.2) | 28.8 (22.0) |
| Alcohol consumption status (%) |  |  |  |
| Never | 3.7 | 4.4 | 5.8 |
| Previous | 2.6 | 3.8 | 6 |
| Current | 93.7 | 91.8 | 88.2 |
| Physical activity (MET-h/week, mean (SD)) | 44.6 (44.2) | 44.8 (45.8) | 40.7 (46.7) |
| Chronotype (%) |  |  |  |
| Morning | 27.6 | 28.2 | 19.4 |
| Morning>evening | 40.2 | 34.4 | 17.6 |
| Evening>morning | 32.2 | 26.7 | 16.3 |
| Evening | 0.0 | 10.6 | 46.7 |
| Duration (h/day, mean (SD)) | 7.2 (0.8) | 7.2 (1.1) | 7.1 (2.0) |
| Daytime napping (%) |  |  |  |
| Never/rarely | 100.0 | 18.2 | 5.1 |
| Sometimes | 0.0 | 73.8 | 74.7 |
| Usually | 0.0 | 8.0 | 20.2 |
| Daytime sleepiness (%) |  |  |  |
| Never/rarely | 89.3 | 68.2 | 44.5 |
| Sometimes | 10.7 | 31.3 | 28.4 |
| Often/always | 0.0 | 0.6 | 27.1 |
| Snoring (%) | 65.9 | 60.5 | 57.4 |
| Insomnia (%) |  |  |  |
| Never/rarely | 27.1 | 22.4 | 19.3 |
| Sometimes | 49.5 | 48.4 | 36.2 |
| Usually | 23.5 | 29.1 | 44.5 |

Abbreviations: BMI: body mass index; GERD: gastro-esophageal reflux disease; MET: Metabolic Equivalent Task; SD: standard deviation.
<sup>a</sup> Composite sleep score was calculated by summing the individual scores of four unhealthy sleep behaviors: evening chronotype, sleep <6 or >9 h/day, daytime napping and usual daytime sleepiness. Each unhealthy sleep behavior was assigned with score 1 otherwise with score 0. The composite sleep score was further categorized into three groups: good (score=0), intermediate (score=1), and poor (score≥2).

### Assessment of covariates

At study enrollment, demographic characteristics and known predictors of esophageal cancer were self-reported, including age, sex, race, education status, smoking status and intensity, alcohol consumption status, and history of gastro-esophageal reflux disease (GERD).^23^ Height was measured (seca 202 stadiometer; seca) to the nearest 0.1 cm, and weight was measured to the nearest 0.1 kg (BC-418 MA body composition analyzer; Tanita Corp). Body mass index (BMI) was derived from weight in kilograms divided by height in meters squared. The Townsend Deprivation Index, a census-based index of material deprivation, was used as a proxy measure for socioeconomic status. Positive values of the index indicate areas with high material deprivation, whereas negative values indicate relative material affluence. Total physical activity in metabolic equivalent task (MET) was derived based on the frequency and duration of walking, and moderate and vigorous activities in the last 7 days, through the International Physical Activity Questionnaire (IPAQ).^24^

### Statistical analyses

Person-years were accrued from the date of initial assessment visit to the date of any cancer diagnoses (excluding non-melanoma skin cancer), the end of follow-up or death, whichever came first. We first examined the associations between each of the six sleep behaviors (chronotype, duration, daytime napping, daytime sleepiness, snoring, and insomnia) and risk of esophageal cancer using age- and multivariable-adjusted Cox proportional hazard models. The covariates included in the multivariable model were established or potential risk factors for esophageal cancer (**eMethods** in the Supplement).

To create a composite sleep score, we adopted the similar approach used previously by only including sleep behaviors with significant impact on esophageal cancer risk.^25–27^ We compared the multivariable models with and without a specific behavior using the likelihood ratio test (LRT). If the *P*_*LRT*_ was ≤0.20, we included this sleep behavior in the composite sleep score. Based on this pre-specified criterion, four sleep behaviors (chronotype, duration, daytime napping, and daytime sleepiness) were selected. We further dichotomized the selected behaviors and assigned a score of 1 for each unhealthy sleep behavior: evening chronotype, sleep <6 or >9 h/day, any daytime napping, and usual daytime sleepiness, and a score of 0 otherwise. We summed the scores across the four behaviors to obtain a composite sleep score ranging from 0 to 4. We further categorized participants into three levels of sleep: good sleep (score=0), intermediate sleep (score=1), and poor sleep (score≥2). Test for linear trend was performed by treating composite sleep score as a continuous variable. Subtype analyses according to histology (EAC *vs*. ESCC) were also performed. For covariates with missing values (0.1-0.8%), we used missing categories.

For EAC, we evaluated whether the association between the composite sleep score and cancer risk differs by genetic predispositions. We tested the interactions using the product terms of continuous composite sleep score and continuous PRS, and *P* for interaction was estimated using the Wald test. We further evaluated the joint associations between composite sleep score and PRS groups (low: Q1-Q2, intermediate: Q3-Q4; high: Q5), using participants with good sleep and low genetic risk as to the reference. In these models, we additionally adjusted for the first ten principal components for ancestry and the genotype array type (UK BiLEVE *vs*. UK Biobank Axiom).^28^

We also conducted multiple sensitivity analyses as described in **eMethods** in the Supplement. All analyses were performed using R (version 4.0.5) and considered significant with 2-sided *P*<0.05.

## Results

Baseline characteristics of the study population are described in **Table 1**. Among 410,428 participants, those with higher composite sleep scores reflecting poorer sleep were more likely to be males, have higher BMI, be current smokers, and with a history of GERD. They were less likely to have a post-college education or be physically active.

During up to 9.3 years of follow-up with 2,799,342 person-years, a total of 410 esophageal cancer cases were diagnosed, with 294 EAC and 95 ESCC. In age-adjusted models, evening chronotype, sleep <6 or >9 h/day, any daytime napping, usual daytime sleepiness, and snoring were associated with an increased risk of esophageal cancer (**Table 2**). After multivariable adjustment, four sleep behaviors had *P*_*LRT*_≤0.2, including evening chronotype, sleep <6 or >9 h/day, any daytime napping, and usual daytime sleepiness. Specifically, compared with individuals with morning chronotype, those who adhere to evening chronotype had a 38% (HR=1.38, 95%CI: 0.98, 1.92) increased risk of esophageal cancer. Participants who slept <6 or >9 h/day had 1.49 (HR=1.49, 95%CI: 1.01, 2.20) and 2.05 (HR=2.05, 95%CI: 1.27, 3.29) times risk of esophageal cancer, compared with individuals who slept 7 hours. Participants who took daytime napping sometimes or usually had 23% (HR=1.23, 95%CI: 1.00, 1.52) and 38% (HR=1.38, 95%CI: 0.98, 1.93) increased esophageal cancer risk, compared with those who never took daytime napping. Compared with those who never had daytime sleepiness, participants with usual daytime sleepiness had a 32% (HR=1.32, 95%CI: 0.83, 2.08) elevated esophageal cancer risk. Snoring and insomnia were not associated with esophageal cancer risk. The results were similar when esophageal cancer diagnosed within the first two years of follow-up were excluded from the analyses (**eTable 2** in the Supplement).

**Table 2.** Sleep behaviors and risk of esophageal cancer, UK biobank 2006-2016.

|  | **N events** | **HR (95%CI) <sup>a</sup>** | **HR (95%CI) <sup>b</sup>** |
| --- | --- | --- | --- |
| Chronotype |  |  |  |
| Morning | 112 | 1 (reference) | 1 (reference) |
| More morning than evening | 136 | 0.97 (0.75, 1.24) | 0.97 (0.76, 1.25) |
| More evening than morning | 111 | 1.06 (0.82, 1.38) | 0.98 (0.75, 1.27) |
| Evening | 51 | 1.68 (1.21, 2.34) | 1.38 (0.98, 1.92) |
| Duration (h/day) |  |  |  |
| <6 | 32 | 1.60 (1.09, 2.35) | 1.49 (1.01, 2.20) |
| 6 | 82 | 1.18 (0.90, 1.55) | 1.13 (0.86, 1.48) |
| 7 | 139 | 1 (reference) | 1 (reference) |
| 8 | 109 | 0.94 (0.73, 1.20) | 0.93 (0.73, 1.20) |
| 9 | 28 | 1.05 (0.70, 1.58) | 0.95 (0.63, 1.43) |
| >9 | 20 | 2.58 (1.61, 4.13) | 2.05 (1.27, 3.29) |
| Daytime napping |  |  |  |
| Never/rarely | 165 | 1 (reference) | 1 (reference) |
| Sometimes | 200 | 1.51 (1.22, 1.85) | 1.23 (1.00, 1.52) |
| Usually | 45 | 2.16 (1.55, 3.01) | 1.38 (0.98, 1.93) |
| Daytime sleepiness |  |  |  |
| Never/rarely | 305 | 1 (reference) | 1 (reference) |
| Sometimes | 85 | 0.84 (0.66, 1.06) | 0.79 (0.62, 1.01) |
| Usually | 20 | 1.58 (1.01, 2.49) | 1.32 (0.83, 2.08) |
| Snoring |  |  |  |
| No | 225 | 1 (reference) | 1 (reference) |
| Yes | 185 | 1.39 (1.14, 1.68) | 1.06 (0.97, 1.30) |
| Insomnia |  |  |  |
| Never | 89 | 1 (reference) | 1 (reference) |
| Sometimes | 198 | 1.02 (0.79, 1.31) | 1.19 (0.93, 1.53) |
| Often/always | 123 | 1.03 (0.79, 1.36) | 1.17 (0.89, 1.55) |

Abbreviations: BMI: body mass index; CI: confidence interval; GERD: gastro-esophageal reflux disease; HR: hazard ratio; MET: Metabolic Equivalent Task.
<sup>a</sup> Adjusted for age at baseline (in years).
<sup>b</sup> Additionally adjusted for sex (male, female), race (White, Black, other), education(pre-/post-college), the Townsend Deprivation Index (in quartiles), history of GERD (yes, no), BMI (underweight, normal, overweight, obese), smoking status and intensity (never smoker, past smoker 1-19 pack-years, past smoker >19 pack-years, past smoker unknown pack-year, current smoker 1-19 pack-years, current smoker >19 pack-years, current smoker with unknown pack-year), and alcohol consumption status (never, previous, current), and physical activity (MET-h/week, in quartiles).

Based on the composite sleep score, compared with participants with a good sleep (score=0), those with an intermediate sleep (score=1) had a 41% (HR=1.41, 95%CI: 1.13, 1.77) increased risk of esophageal cancer, and individuals with a poor sleep (score ≥ 2) had a 79% increased risk (HR=1.79, 95%CI: 1.32, 2.42) (*P*_*trend*_<0.001). These findings were similar in sensitivity analyses that excluded esophageal cancer diagnosed within 2 years after baseline (**eTable 3** in the Supplement), with finer risk score groups (**eTable 4** in the Supplement), when participants with a history of GERD were removed from the analysis (**eTable 5** in the Supplement), or stratified by BMI categories (<25 and ≥25 kg/m^2^, **eTable 6** in the Supplement).

**Table 3.** Composite sleep score and risk of esophageal cancer overall and according to histology.

| Composite sleep score <sup>a</sup> | Esophageal cancer |  | EAC |  | ESCC |  |
| --- | --- | --- | --- | --- | --- | --- |
|  | *N* events | HR (95%CI) <sup>b</sup> | *N* events | HR (95%CI) <sup>b</sup> | *N* events | HR (95%CI) <sup>b</sup> |
| Good (score=0) | 123 | 1 (reference) | 84 | 1 (reference) | 35 | 1 (reference) |
| Intermediate (score=1) | 219 | 1.41 (1.13, 1.77) | 157 | 1.38 (1.05, 1.80) | 50 | 1.38 (0.89, 2.14) |
| Poor (score ≥2) | 68 | 1.79 (1.32, 2.42) | 53 | 1.88 (1.32, 2.67) | 10 | 1.16 (0.56, 2.38) |
| *P* trend <sup>c</sup> |  | <0.001 |  | <0.001 |  | 0.340 |

Abbreviations: CI: confidence interval; EAC: esophageal adenocarcinoma; ESCC: esophageal squamous cell carcinoma; HR: hazard ratio.
<sup>a</sup> Composite sleep score was calculated by summing the individual scores of four unhealthy sleep behaviors: evening chronotype, sleep <6 or >9 h/day, daytime napping and usual daytime sleepiness. Each unhealthy sleep behavior was assigned with score 1 otherwise with score 0. The composite sleep score was further categorized into three groups: good (score=0), intermediate (score=1), and poor (score≥2).
<sup>b</sup> Adjusted for the same set of covariates as model b in Table 2.
<sup>c</sup> *P* value for trend was calculated using the composite sleep score as a continuous variable (i.e., 0-4).

We also evaluated the associations according to the histology of esophageal cancer. The elevated risk between the composite sleep score and esophageal cancer was largely driven by EAC (*P*_*trend*_<0.001) but not ESCC (*P*_*trend*_=0.340) (**Table 3**). Compared with participants with a good sleep, those with an intermediate or poor sleep had 38% (HR=1.38, 95%CI: 1.05, 1.80) and 88% (HR=1.88, 95%CI: 1.32, 2.67) increased risks of EAC, respectively.

For EAC, we further examined the interaction between the composite sleep score and PRS as well as their joint associations. We observed similar associations between the composite sleep score and EAC risk within participants with low (Q1-Q2 of PRS), intermediate (Q3-Q4), or high genetic risk (Q5) (**Figure 1**), respectively. No interactions were observed (*P*_*interaction*_=0.791) between genetic risk and sleep behaviors for EAC risk. In the joint analyses, compared to participants with a good sleep and low genetic risk, those with a poor sleep and high genetic risk had an HR of 6.64 (95%CI: 3.10, 14.20) (**Figure 1, eTable 7** in the Supplement).

## Discussion

In a large prospective cohort, we examined the associations between major sleep behaviors and their collective impact on esophageal cancer risk. We also assessed the associations according to histology and genetic susceptibilities. Overall, evening chronotype, short or long duration (<6 or >9 h/day), daytime napping, and usual daytime sleepiness were associated with increased esophageal cancer risk. The elevated risks further increased with a greater number of the above-mentioned high-risk behaviors. These positive associations were driven primarily by EAC, regardless of their genetic risk.

Growing evidence has linked unhealthy sleep behaviors with increased risk of multiple cancers,^29–32^ yet studies on esophageal cancer are sparse and largely limited to single sleep trait such as duration, snoring,^8^ and daytime napping.^33^ One case-control study from China reported that short sleep (<7 h/day) was associated with an increased risk of ESCC.^8^ A Mendelian randomization study among UK Biobank participants showed that genetic liability to short (<7 h/day) or long sleep (≥9 h/day) duration was not causally linked with esophageal cancer risk,^34^ yet SNPs associated with extreme short (<6 h/day) or long sleep (>9 h/day) have not been examined. Our findings that extreme short or long sleep were both associated with increased risk of esophageal cancer indicate the further need to evaluate extreme duration of sleep. Indeed, individuals who sleep 5-6 hours were found to have a higher risk of Barrett’s esophagus, compared with those who sleep 7-8 hours each day.^7^ For daytime napping, the UK Million Women Study reported no association between sometimes/usually *vs*. rare/never daytime napping with risk of esophageal cancer after an average of 7.4 years of follow up,^33^ while our analyses supported a positive association with a longer follow up and even in sensitivity analyses that excluded the first two years of esophageal cancer diagnoses to minimize the impact of undiagnosed cancers. In addition to reporting probable associations with individual sleep traits, our analyses for the first time constructed a composite sleep score to capture collective impact of sleep behaviors. Besides reporting a linear relationship between the number of unhealthy sleep behaviors and the risk of esophageal cancer, we also showed that these positive associations were driven by EAC and independent of genetic risk, though the possibility for ESCC could not be fully ruled out due to its limited number of cases. Taken together, our findings suggest that unhealthy sleep behaviors are likely important for EAC etiology and validations are warranted.

The biological mechanisms linking unhealthy sleep behaviors and the risk of esophageal cancer remain to be elucidated. Evening chronotype and shortened sleep are linked with disrupted circadian rhythm, causing alternations in hormone and metabolic profiles, which are risk factors for esophageal cancer.^35^ Additionally, melatonin, which is associated with lower risk of EAC in animal trials, may be suppressed for secretion among participants with evening chronotype.^36^ Disruption of circadian physiology owing to shortened or prolonged sleep and daytime sleepiness could potentially result in gastrointestinal diseases such as GERD,^37^ a well-established risk factor for EAC.^11^ Having a history of GERD with excess sleep could lead to a prolonged period of esophageal acid exposure to more proximal regions of the esophagus, which may further increase the risk for Barrett’s esophagus and EAC.^38^ In addition, a recent Mendelian randomization study revealed that daytime napping is causally linked with greater waist circumference and high blood pressure,^15^ which are risk factors for EAC^39^ and ESCC.^40^ Excessive daytime sleepiness occurs frequently among people with narcolepsy who tend to have autoimmune disorder,^41^ a risk factor for cancer.^42^ For example, narcolepsy patients are shown to have higher level of human leukocyte antigen (HLA) DRB1*1501^43^ and HLA DQB1*0602,^43,44^ resulting in a higher gastric cancer susceptibility and esophageal cancer risk.^45^ Intriguingly, GWAS studies have identified the association of MTRNR2L9 with EAC as well as chronotype, sleep duration, daytime napping, daytime sleepiness, insomnia, and snoring, suggesting a potential genetic pathway between sleep traits and EAC risk.^13,17,19^

Primary strengths of our study include a large sample size and a prospective study design. More notably, our study is among the first to comprehensively examine the associations of sleep behaviors with esophageal cancer risk through integrating multiple sleep behaviors. We also evaluated the interaction and joint association between sleep behaviors and EAC genetic susceptibility. Our study has a few limitations. First, all sleep behaviors were self-reported. These subjective measures on sleep behaviors might not accurately reflect their sleep condition. However, non-differential misclassifications will likely bias the study association toward the null. Furthermore, Mendelian randomization studies have validated these traits against accelerometers and confirmed their causal links breast cancer^34^ and lung cancer.^46^ In addition, residual confounding from unknown or unmeasured factors may still exist despite our efforts in adjusting for major confounders. Lastly, findings were based on the UK Biobank participants composed of mostly European descendants and may not generalize to other populations.

## Conclusions

Unhealthy sleep behaviors, including evening chronotype, sleep <6 or >9 h/day, daytime napping, and usual daytime sleepiness, were associated with an increased risk of esophageal cancer, primarily EAC, independently of genetic risk. These findings suggest that sleep behaviors may serve as modifiable factors for the prevention of esophageal cancer, particularly EAC.

## Supporting information

eMethods, eTables and eFigures

## Data Availability

All data produced are available at UK Biobank.

## Author Contributions

Wang, Tian and Cao had full access to all the data in the study and take responsibility for the integrity of the data and the accuracy of the data analysis.

Concept and design: Wang, Cao.

Acquisition, analysis, or interpretation of data: Wang, Tian, Zong, Jeon, Luo, Cao.

Drafting of the manuscript: Wang, Tian, Cao

Critical revision of the manuscript for important intellectual content: All authors.

Statistical analysis: Wang, Tian, Zong.

Obtained funding: Cao.

Administrative, technical, or material support: Cao.

Supervision: Cao.

