## Supplementary material for "Association of Sleep Behaviors with Risk of Esophageal Cancer": eMethods, eTables and eFigures

### eMethods. Statistical Analysis

#### Covariates in multivariable model

The covariates included in the multivariable models were age (in years), sex (female or male), race (White, Black, or other), education status (pre- or post-college), Townsend Deprivation Index (in quartiles), history of gastro-esophageal reflux disease (GERD, yes or no), body mass index (BMI, underweight: <18.5 kg/m2, normal: 18.5-24.9 kg/m2, overweight: 25-29.9 kg/m2, obese: ≥30 kg/m2), smoking status and intensity (never smoker, past smoker 1-19 pack-years, past smoker >19 pack-years, past smoker unknown pack-year, current smoker 1-19 pack-years, current smoker >19 pack-years, current smoker unknown pack-year), alcohol consumption status (never, past, or current), and total physical activity (metabolic equivalent task [MET] h/week, in quartiles).

#### Exclusion criteria for polygenic risk score (PRS) related analyses

Details of the genotyping process in UK Biobank were described elsewhere.[1,2] We acquired imputed data of 487,409 participants initially. Using UK Biobank provided quality control sample file, participants were excluded if they were outliers with high missingness or for which heterozygosity rates were not explained by runs of homozygosity analysis nor mixed ethnicity, or were marked as sex chromosome aneuploidy.[2,3] The UK Biobank dataset estimated a kinship coefficient for each pair of samples using KING's robust estimator.[4] We further excluded third-degree (or higher) related individuals (kinship coefficient ≥ 0.0442).[5] After these exclusions, a total of 278,893 participants remained in the analyses.

#### Sensitivity analyses

The following sensitivity analyses were conducted : 1) excluded participants who were diagnosed with esophageal cancer within the first two years of follow-up to reduce the likelihood of reverse causation, i.e., having poor sleep behaviors due to undiagnosed esophageal cancer at baseline, 2) expanded the categories of composite sleep score to four groups with sleep score as 0, 1, 2 and ≥3 to explore the dose-response relationship, 3) excluded individuals with a history of GERD to examine the independent association not mediated by GERD, and 4) stratified by BMI categories (<25 and ≥25 kg/m2) to assess whether the associations was similarly observed within normal weight vs. overweight/obese participants.

**eTable 1. Association between individual SNPs (previously associated with EAC or Barrett's esophagus) and risk of EAC.**

| SNP | Position | *Gene* | Published OR for risk allele[6–8] | Risk allele frequency in UK Biobank | OR in UK Biobank |
|---|---|---|---|---|---|
| rs7255 | 2:20878820 | *GDF7-LDAH* | 1.17 | 37% (T) | 1.02 |
| *rs13397172* | 2:200045039 | *SATB2* | 1.13 | 45% (C) | 1.06 |
| rs3072 | 2:20878406 | *GDF7* | 1.14 | 29% (C) | 1.12 |
| rs2687202 | 3:70929983 | *FOXP1* | 1.13 | 24% (T) | 1.21 |
| rs9823696 | 3:183783353 | *HTR3C* | 1.17 | 30% (A) | 1.14 |
| rs62423175 | 6:62195368 | *KHDRBS2-MTRNR2L9* | 1.23 | 14% (A) | 1.22 |
| rs17451754 | 7:117256712 | *CFTR* | 1.25 | 71% (G) | 1.00 |
| rs2188554 | 7:117040117 | *ASZ1* | 1.23 | 66% (A) | 1.12 |
| rs10108511 | 8:11435516 | *LINC00208- BLK* | 1.12 | 38% (T) | 1.25 |
| rs17749155 | 8:10068073 | *MSRA* | 1.14 | 13% (A) | 1.19 |
| rs7852462 | 9:100310501 | *TMOD1* | 1.08 | 49% (C) | 1.18 |
| rs11789015 | 9:96716028 | *BARX1* | 1.20 | 59% (A) | 1.51 |
| rs1247942 | 12:114673723 | *LOC1053699 96-TBX5* | 1.11 | 48% (G) | 0.96 |
| rs2464469 | 15:58362025 | *ALDH1A2* | 1.11 | 34% (G) | 1.22 |
| rs1979654 | 16:86396835 | *LOC732275-FOXF1* | 1.11 | 34% (G) | 0.92 |
| rs10419226 | 19:18803172 | *CRTC1* | 1.18 | 37% (T) | 1.06 |
| rs10423674 | 19:18817903 | *CRTC1* | 1.19 | 54% (C) | 1.14 |

Abbreviations: EAC: esophageal adenocarcinoma; OR: odds ratio; SNP: single nucleotide polymorphism.

**eTable2. Sleep behaviors and risk of esophageal cancer diagnosed after 2 years of enrollment.**

| | *N* events | HR (95%CI) [a] | HR (95%CI) [b] |
|---|---|---|---|
| Chronotype | | | |
| Morning | 79 | 1 (reference) | 1 (reference) |
| More morning than evening | 112 | 1.13 (0.84, 1.50) | 1.14 (0.86, 1.53) |
| More evening than morning | 79 | 1.07 (0.78, 1.46) | 0.99 (0.73, 1.36) |
| Evening | 44 | 2.05 (1.41, 2.96) | 1.72 (1.18, 2.50) |
| Duration (h/day) | | | |
| <6 | 23 | 1.42 (0.91, 2.22) | 1.32 (0.84, 2.08) |
| 6 | 64 | 1.13 (0.83, 1.54) | 1.08 (0.79, 1.47) |
| 7 | 113 | 1 (reference) | 1 (reference) |
| 8 | 81 | 0.86 (0.65, 1.14) | 0.86 (0.64, 1.14) |
| 9 | 16 | 0.75 (0.44, 1.26) | 0.68 (0.40, 1.15) |
| >9 | 17 | 2.74 (1.65, 4.57) | 2.20 (1.31, 3.69) |
| Daytime napping | | | |
| Never/rarely | 130 | 1 (reference) | 1 (reference) |
| Sometimes | 151 | 1.45 (1.14, 1.84) | 1.19 (0.94, 1.51) |
| Usually | 33 | 2.04 (1.39, 3.00) | 1.31 (0.89, 1.94) |
| Daytime sleepiness | | | |
| Never/rarely | 235 | 1 (reference) | 1 (reference) |
| Sometimes | 65 | 0.84 (0.65, 1.10) | 0.79 (0.60, 1.05) |
| Usually | 14 | 1.45 (0.85, 2.49) | 1.21 (0.70, 2.09) |
| Snoring | | | |
| No | 175 | 1 (reference) | 1 (reference) |
| Yes | 139 | 1.34 (1.07, 1.67) | 1.01 (0.81, 1.28) |
| Insomnia | | | |
| Never | 69 | 1 (reference) | 1 (reference) |
| Sometimes | 150 | 1.00 (0.75, 1.33) | 1.17 (0.88, 1.55) |
| Often/always | 95 | 1.03 (0.79, 1.36) | 1.18 (0.86, 1.61) |

Abbreviations: CI: confidence interval; GERD: gastro-esophageal reflux disease; HR: hazard ratio.
[a] Adjusted for age at baseline (in years).
[b] Adjusted for the same set of covariates as model b in Table 2.

**eTable 3. Composite sleep score and risk of esophageal cancer diagnosed after 2 years of enrollment.**

| Composite sleep score [a] | Esophageal cancer | | EAC | | ESCC | |
|---|---|---|---|---|---|---|
| | *N* events | HR (95%CI) [b] | *N* events | HR (95%CI) [b] | *N* events | HR (95%CI) [b] |
| Good (score=0) | 95 | 1 (reference) | 67 | 1 (reference) | 25 | 1 (reference) |
| Intermediate (score=1) | 166 | 1.41 (1.09, 1.82) | 122 | 1.38 (1.02, 1.86) | 38 | 1.48 (0.89, 2.47) |
| Poor (score ≥2) | 53 | 1.85 (1.31, 2.62) | 39 | 1.79 (1.20, 2.69) | 9 | 1.52 (0.70, 3.32) |
| *P* trend [c] | | <0.001 | | 0.002 | | 0.151 |

Abbreviations: CI: confidence interval; EAC: esophageal adenocarcinoma; ESCC: esophageal squamous cell carcinoma; HR: hazard ratio.

[a] Composite sleep score was calculated by summing the individual scores of four unhealthy sleep behaviors: evening chronotype, sleep <6 or >9 h/day, daytime napping and usual daytime sleepiness. Each unhealthy sleep behavior was assigned with score 1 otherwise with score 0. The composite sleep score was further categorized into three groups: good (score=0), intermediate (score=1), and poor (score≥2).

[b] Adjusted for the same set of covariates as model b in Table 2.

[c] *P* value for trend was calculated using composite sleep score as a continuous variable (i.e., 0-4).

**eTable 4. Composite sleep score with finer categories and risk of esophageal cancer and adenocarcinoma.**

| Composite sleep score [a] | Esophageal cancer | | EAC | |
|---|---|---|---|---|
| | *N* events | HR (95%CI) [b] | *N* events | HR (95%CI) [b] |
| 0 | 123 | 1 (reference) | 84 | 1 (reference) |
| 1 | 219 | 1.41 (1.13, 1.77) | 157 | 1.38 (1.05, 1.81) |
| 2 | 57 | 1.71 (1.24, 2.36) | 45 | 1.82 (1.26, 2.64) |
| 3-4 | 11 | 2.34 (1.25, 4.38) | 8 | 2.28 (1.10, 4.76) |
| *P* trend [c] | | <0.001 | | <0.001 |

Abbreviations: CI: confidence interval; EAC: esophageal adenocarcinoma; HR: hazard ratio.

[a] Composite sleep score was calculated by summing the individual scores of four unhealthy sleep behaviors: evening chronotype, sleep <6 or >9 h/day, daytime napping and usual daytime sleepiness. Each unhealthy sleep behavior was assigned with score 1 otherwise with score 0. The composite sleep score was further categorized into four groups: score of 0, 1, 2, and 3-4.

[b] Adjusted for the same set of covariates as model b in Table 2.

[c] *P* value for trend was calculated using composite sleep score as a continuous variable.

#### eTable 5. Composite sleep score and risk of esophageal cancer excluding participants with history of GERD

| Composite sleep score [a] | Esophageal cancer | | EAC | |
|---|---|---|---|---|
| | ***N* events** | **HR (95%CI) [b]** | ***N* events** | **HR (95%CI) [b]** |
| Good (score=0) | 114 | 1 (reference) | 69 | 1 (reference) |
| Intermediate (score=1) | 209 | 1.47 (1.16, 1.85) | 147 | 1.48 (1.12, 1.95) |
| Poor (score ≥2) | 64 | 1.85 (1.35, 2.53) | 47 | 2.00 (1.39, 2.88) |
| *P* trend [c] | | <0.001 | | <0.001 |

Abbreviations: CI: confidence interval; EAC: esophageal adenocarcinoma; GERD: gastro-esophageal reflux disease; HR: hazard ratio.

[a] Composite sleep score was calculated by summing the individual scores of four unhealthy sleep behaviors: evening chronotype, sleep <6 or >9 h/day, daytime napping and usual daytime sleepiness. Each unhealthy sleep behavior was assigned with score 1 otherwise with score 0. The composite sleep score was further categorized into three groups: good (score=0), intermediate (score=1), and poor (score≥2).

[b] Adjusted for the same set of covariates as model b in Table 2 excluding history of GERD.

[c] *P* value for trend was calculated using composite sleep score as a continuous variable.

#### eTable 6. Composite sleep score and risk of esophageal cancer by BMI group (<25 and ≥25)

| Composite sleep score [a] | Esophageal cancer | | EAC | | ESCC | |
|---|---|---|---|---|---|---|
| | *N* events | HR (95%CI) [b] | *N* events | HR (95%CI) [b] | *N* events | HR (95%CI) [b] |
| **BMI < 25** kg/m$^2$ | | | | | | |
| Good (score=0) | 33 | 1 (reference) | 13 | 1 (reference) | 18 | 1 (reference) |
| Intermediate (score=1) | 51 | 1.61 (1.03, 2.52) | 25 | 1.79 (0.91, 3.53) | 24 | 1.53 (0.82, 2.84) |
| Poor (score ≥2) | 12 | 1.74 (0.88, 3.42) | 8 | 2.56 (1.03, 6.32) | 2 | 0.61 (0.14, 2.68) |
| *P* trend [c] | | 0.034 | | 0.029 | | 0.725 |
| **BMI ≥ 25** kg/m$^2$ | | | | | | |
| Good (score=0) | 90 | 1 (reference) | 71 | 1 (reference) | 17 | 1 (reference) |
| Intermediate (score=1) | 167 | 1.35 (1.04, 1.76) | 132 | 1.31 (0.98, 1.76) | 25 | 1.23 (0.66, 2.30) |
| Poor (score ≥2) | 55 | 1.79 (1.27, 2.52) | 44 | 1.75 (1.19, 2.57) | 8 | 1.58 (0.67, 3.73) |
| *P* trend [c] | | <0.001 | | 0.004 | | 0.292 |

Abbreviations: CI: confidence interval; EAC: esophageal adenocarcinoma; ESCC: esophageal squamous cell carcinoma; HR: hazard ratio.

[a] Composite sleep score was calculated by summing the individual scores of four unhealthy sleep behaviors: evening chronotype, sleep <6 or >9 h/day, daytime napping and usual daytime sleepiness. Each unhealthy sleep behavior was assigned with score 1 otherwise with score 0. The composite sleep score was further categorized into three groups: good (score=0), intermediate (score=1), and poor (score≥2).

[b] Adjusted for the same set of covariates as model b in Table 2.

[c] *P* value for trend was calculated using composite sleep score as a continuous variable.

**eTable 7. Joint associations of composite sleep score and genetic risk with subsequent risk of EAC.**

| Genetic risk [b] | Composite sleep score [a] | | |
|---|---|---|---|
| | Good (score=0) | Intermediate (score=1) | Poor (score ≥2) |
| **Low (Q1-Q2)** | | | |
| *N* events | 13 | 38 | 9 |
| HR (95%CI) [c] | 1 (Reference) | 2.16 (1.15, 4.06) | 2.01 (0.85, 4.73) |
| **Intermediate (Q3-Q4)** | | | |
| *N* events | 27 | 39 | 15 |
| HR (95%CI) [c] | 2.03 (1.05, 3.94) | 2.16 (1.15, 4.05) | 3.52 (1.66, 7.44) |
| **High (Q5)** | | | |
| *N* events | 17 | 30 | 14 |
| HR (95%CI) [c] | 2.56 (1.25, 5.28) | 3.27 (1.70, 6.28) | 6.64 (3.10, 14.20) |

Abbreviations: CI: confidence interval; EAC: Esophageal adenocarcinoma; HR: hazard ratio.

[a] Composite sleep score was calculated by summing the individual scores of four unhealthy sleep behaviors: evening chronotype, sleep <6 or >9 h/day, daytime napping and usual daytime sleepiness. Each unhealthy sleep behavior was assigned with score 1 otherwise with score 0. The composite sleep score was further categorized into three groups: good (score=0), intermediate (score=1), and poor (score≥2).

[b] Genetic risk was defined by quintiles of PRS: low-(Q1-Q2), intermediate- (Q3-Q4), and high-risk (Q5).

[c] Adjusted for the same set of covariates as model b in Table 2, the first ten principal components for ancestry, and genotype array type.

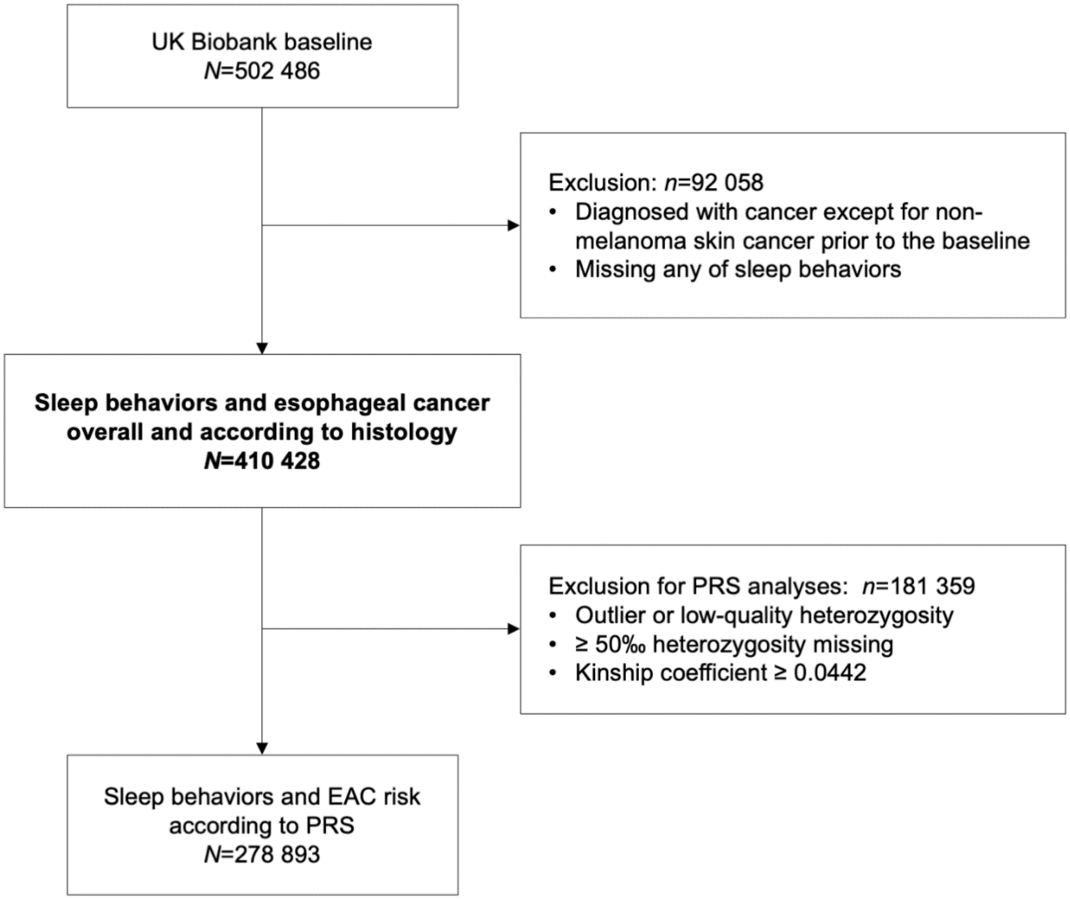


**eFigure 1. Flow chart of study population**

Abbreviations: EAC: esophageal adenocarcinoma; PRS: polygenic risk score.
